# From Concept to Reality: Examining India’s Clinical Decision Support System (CDSS) Challenges & Opportunities

**DOI:** 10.1101/2023.04.02.23288046

**Authors:** Aswini Misro, Anushka Mehta, Paul Whittington, Huseyin Dogan, Nishikant Mishra, Naim Kadoglou, Selva Theivacumar

## Abstract

**Background:** Clinical Decision Support Systems (CDSS) are rapidly altering the face of healthcare and their potential to improve patient outcomes has been exploited, in some countries. This study aims to explore the current landscape of the Indian healthcare sector to identify the favourability of current practises, organisational and infrastructural readiness, attitudes and concerns of the stakeholders concerning the implementation of CDSS.

**Methods:** The methodology that this study used was carrying out structured interviews comprising of 16 close-ended questions, split into three sub-categories. There was a total of 61 interviews were conducted with medical and administrative staff in public and privately run facilities, present in Tier 1 and Tier 2 cities in India. The study will focus on hospitals in Tier 1 cities as these are in a position to bring technological transformation.

**Results:** The results identified various trends and patterns that would likely govern the incorporation of CDSS. A large proportion of the experts answered positively about the current level of digitalisation of their workplace and the availability of funds for future innovation, indicating high favourability for CDSS. Various roadblocks were isolated with respect to stakeholder attitudes, standardisation of care and general knowledge about CDSS and that in two cities, privately owned facilities were better equipped than state-run facilities.

**Conclusions:** There have been many recent initiatives in India to promote digital health. Performing a CDSS cost-effectiveness study will demonstrate the benefits of using CDSS in the country to overcome any adoption hesitancies.

## 1.0 Background

With growing inequalities, an enormous population and a vast talent pool, the Indian healthcare sector is prime for the successful implementation of sustainable and innovative technologies like Clinical Decision Support Systems (CDSS). CDSS is a component of Health Information Technology (Health IT) which tries to incorporate Artificial Intelligence (AI) to improve patient outcomes. It is based on the principle of developing and applying algorithms to clinical data and formulating advanced diagnostic assessments, risk predictions, prognostications and treatment strategies, aimed at ultimately improving overall healthcare.

The Indian healthcare system has been ranked 112 out of 190 countries in the World Health Report published by WHO^1^ in 2000 (1). However, India has a competitive edge over some of the better-ranked economies due to the high percentage of skilled professionals, the size of the economy, and the cost-efficient medical and surgical treatments offered. The Indian healthcare sector is an integral contributor to the nation’s revenue and employment. The public and private healthcare delivery systems are the two major categories of the Indian healthcare delivery systems. While the public healthcare system is streamlined to address the basic healthcare needs specifically of the rural population, the private care system caters to the more advanced medical needs with several secondary, tertiary, and quaternary private institutions concentrated in top-tier cities (2).

Prehistorically, several forms of traditional medicine were popular in India, some of which still exist. The advent of modern medicine in the subcontinent can be traced back to the 17^th^ century (3). Currently, the doctor-to-patient ratio in India is 1: 1456 against the WHO recommendation of 1:1000 (4). Despite rapid growths and wide expansions in the healthcare arena in recent years, most of the nation’s healthcare is still predigital, written case sheets and radiological films are still more common than their electronic counterparts. Health IT has revolutionised the way medicine is practised, over the globe. In the last decade, the country has seen unprecedented booms in technology. The push for digitalisation has seen more institutions incorporate basic technologies in their healthcare systems. Currently, EHRs^2^, online appointment scheduling, and e-commerce for home delivery of medicines are gaining popularity. With digitalisation, the urge for standardisation of care through the development of standard treatment guidelines has garnered due attention (5).

The main role of CDSS is to assist clinicians rather than make decisions for clinicians, with a better analysis of individual ‘patients’ needs (6). The healthcare industry which is deemed to be one of the most complex and dynamic sectors in India is expected to reach a staggering $372 billion by 2022. The hospital industry which comprises 80% of the sector is growing at a compound annual growth rate of 16-17% and is predicted to hit $132 billion by 2023 (7). According to the Bank for International Settlements (BIS)^3^ research report published in 2018, the Indian CDSS market was valued at $43.8 million. The global and Indian CDSS markets were projected to reach $10.83 billion and $206.1 million, respectively, by 2025. The annual cost savings due to the incorporation of AI in healthcare is predicted to be $150 billion, by 2026 (8). Given the positive trend and outlook of the CDSS market, we studied the Indian market readiness for adopting such an innovative digital product in its highly regulated healthcare environment.

The research question to be addressed in this study is; “Are the Indian healthcare institutions ready to incorporate a CDSS into clinical practice?”‘. The findings obtained from the study will contribute to answering this question.

A few studies have been conducted on the applicability of CDSS in rural India, particularly for use by public non-physician healthcare workers, and physicians working in resource-poor settings in the domain of cardiovascular health and psychiatry. There have been many positive outcomes in regard to the use of CDSS. However, there is not enough quality data available regarding its adoption in any setting other than the primary care environment. The current study is, therefore, focused on the application of CDSS in secondary and tertiary care settings, especially in the domain of cancer care (9-11).

## 2.0 Methods

### 2.1 Aim

The aim of the study is to critically analyse clinical care at secondary care and tertiary care hospitals in India, to gain an insight into the concerns and attitudes towards the incorporation of CDSS into patient practises, which are crucial determinants of the future scope of CDSS in the country (12).

### 2.2 Study Design

This study followed a semi-structured interview design where the participants were asked a set of closed-ended pre-determined questions (e.g., “Are you ready to take the initiative and leadership to bring the change in your organisation if CDSS is given free of charge?”) followed by an open-ended exploratory question (e.g., “How do you view the present and future of CDSS in your personal or institutional clinical practice?”) that would reflect their knowledge, awareness and attitudes towards CDSS, while also provide useful information about the current healthcare practises. All the interviews followed the same prescribed structure. All the interviews were conducted face-to-face in the participants’ offices, over a three-month period. The language used for the face-to-face interviews was English. None of the interviews was repeated. The interviewer was provided with the sequence of the interview e.g. structured questionnaire to start with followed by an open-ended question to explore the circumstances further. The participants were asked a total of 16 questions which were grouped into 3 categories; current healthcare practises, organisational & infrastructural limitations, and personal attitudes towards CDSS. An interview guide was developed with the subcategories which were used in this study. The purpose of the semi-structured interview is to give structure to the entire interview while generating a sufficient amount of quality data that allows comparison across the hospitals and regions. The provision of subjective opinion promotes flexibility and inclusivity to capture the motive, and emotions of the interviewee. Some example questions from each category are, ‘Do you have electronic health records for the clinical services?’, ‘Do you know or have heard of the Clinical Decision Support System?’ and ‘Do you think CDSS can add value to your existing clinical practices?’. The complete set of questions is provided in Appendix 1. The participants could answer either yes, no or cannot say. They were also encouraged to explain their answers and the responses were recorded.

### 2.3 Settings and Study Participants

To maximise the generalisability of the results and to include the perspectives of diverse potential users, the study took the following characteristics into consideration - ownership of the facility, current professional positions held by the participants and the location of the medical institutions that employed the study participants. 25 hospitals were visited in Thiruvananthapuram, Chandigarh and other Tier 1 and 2 cities in India including New Delhi and Mumbai, between the months of August and October 2019. These included corporate, state and central government-run facilities. In India, Tier-1 is defined as being highly developed with good facilities and a population of over 100,000, whereas Tier-2 are in the process of being developed and has a population of between 50,000 and 99,999. The participants of the interviews included medical and admin staff.

The recruitment of participants was carried out using a mixed methodology, e.g., social media (LinkedIn and WhatsApp messaging services) and email. We contacted the administrators of 2 surgical societies in order to recruit the participants necessary for this study. The leaflet containing our survey was posted on two WhatsApp groups of senior surgeons and hospital administrators. Out of the 37 people who responded, 22 were eligible to take part in this survey. The eligibility criteria were that they are a senior consultant (e.g., clinical practice in a cancer care speciality for at least 15 years), senior hospital administrator, or head of nursing or technology. 120 eligible participants were emailed about the survey. Out of those, 18 responded. We also found 22 more participants through social media (e.g., LinkedIn). The study included 14 private institutions and 11 government hospitals from Tier 1 and Tier 2 cities of India. Among those, 13 hospitals were in Tier 1 cities and a further 12 hospitals were from Tier 2 cities. 61 out of 62 experts requested the interview and participated in the study. There was a high participation rate of 98.4 % in this study. This is because 61 out of 62 experts that were requested for the interview took part in this study with one declining.

Table 1 shows that 32 participants recruited for the study were employed in facilities in Tier 1 cities while 29 participants worked in hospitals in Tier 2 cities. 30 and 31 participants from the private and public healthcare delivery systems participated in the study, respectively.

**Table 1:** Service Sectors of Study Participants

| Service Sector | Number of Participants |
| --- | --- |
| Oncology | 18 |
| Medical Services | 6 |
| Senior Medical or Surgical Oncology Consultants | 20 |
| Senior Hospital Administrators | 12 |
| Heads of Nursing | 3 |
| Chiefs of Healthcare Technology | 3 |

User data was collected and analysed using Hall’s 7 Stages of Concern model (13), with 0 being Unconcerned and every subsequent stage of increasing concern e.g. 1 = Informational (learning and weighing risks and benefits), 2 = Personal (perceived potential personal concerns and risks), 3 = Management (concerns related to the management of system), 4 = Consequence (impact on other users and environment), 5 = Collaborative (e.g. concerns on the integrated multi-disciplinary workflow), 6 = Refocusing (e.g. ideas for improvements). Stage-0 concern was for uninformed users, while stage-1 was for users who were uninformed but found it intriguing. Stage-2 concerns were related to those subjects who had personal concerns about the impact on their personal circumstances, such as loss of autonomy and external imposition. The stage-3 concerns included logistics issues related to the integration of CDSS into the clinical environment. The level-4 concerns were related to value generated, return on investment (ROI), risk and threat perception (collective).

### 2.4 Data Collection and Analysis

Data from the 61 interviews was collected from the semi-structured interviews conducted in the hospital settings. Following completion of the pre-determined closed questions of the interviews, the interviewee asked to provide their view on their current working environment and feasibility of CDSS. While the responses of the pre-determined structured questions were recorded on a paper-based note, the entire interview was recorded using microphone with the user’s consent and they were later transcribed for the purpose of future referencing. The data collected was analysed and recurrent themes were identified. The responses were grouped into the 3 broad categories that was pre-determined. Comparisons were then made among the sub-groups, with respect to the demographic differences among the study participants. However, the qualifications and the experiences of the individual participants were not considered for analysis.

## 3.0 Results

Out of 61 interviewees, there were 18 oncology professors, 6 directors of medical services, 10 senior consultants in medical oncology, 9 senior consultants in surgical oncology, 4 medical superintendent and 8 senior hospital administrators. The mean age of the participants was 52.2 years and the age distribution is shown in Fig. 1. There are 50 male and 11 female participants in total. While the responses were directed to the adoption of CDSS, they also reflected the infrastructural, people, and process readiness needed for its adoption.

**Fig. 1:**
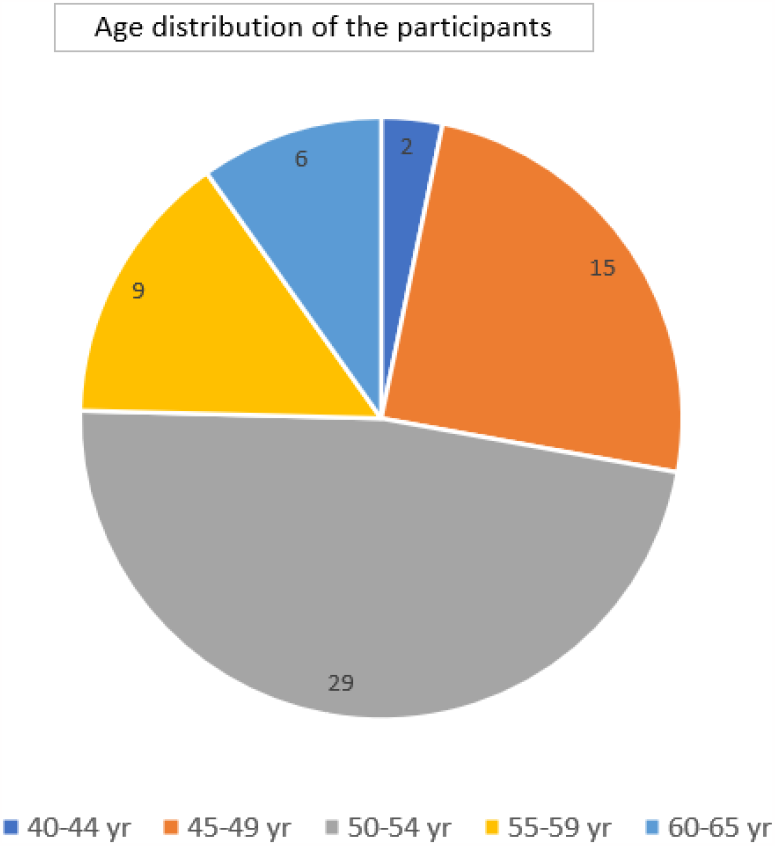
The age distribution of the interviewed participants. Out of total of 976 responses from 61 participants (e.g., 16 sets of responses from 61 participants), 227 were favourable to the adoption of CDSS into clinical practice while 571 were unfavourable. There was a total of 128 “don’t know” responses. Out of the favourable responses, 61.2% were from Tier-1 cities while 38.8% were from Tier-2 cities. In the user adoption issues category, an overwhelming 97% of the favourable responses were from Tier-1 cities while 3% were from Tier-2 cities.

### 3.1 Analysis of the favourability of current practises for CDSS integration

On questions posed to analyse the favourability of current practises for CDSS integration, the majority of answers were not in favour of AI implementation, see Fig. 2.

**Fig. 2.**
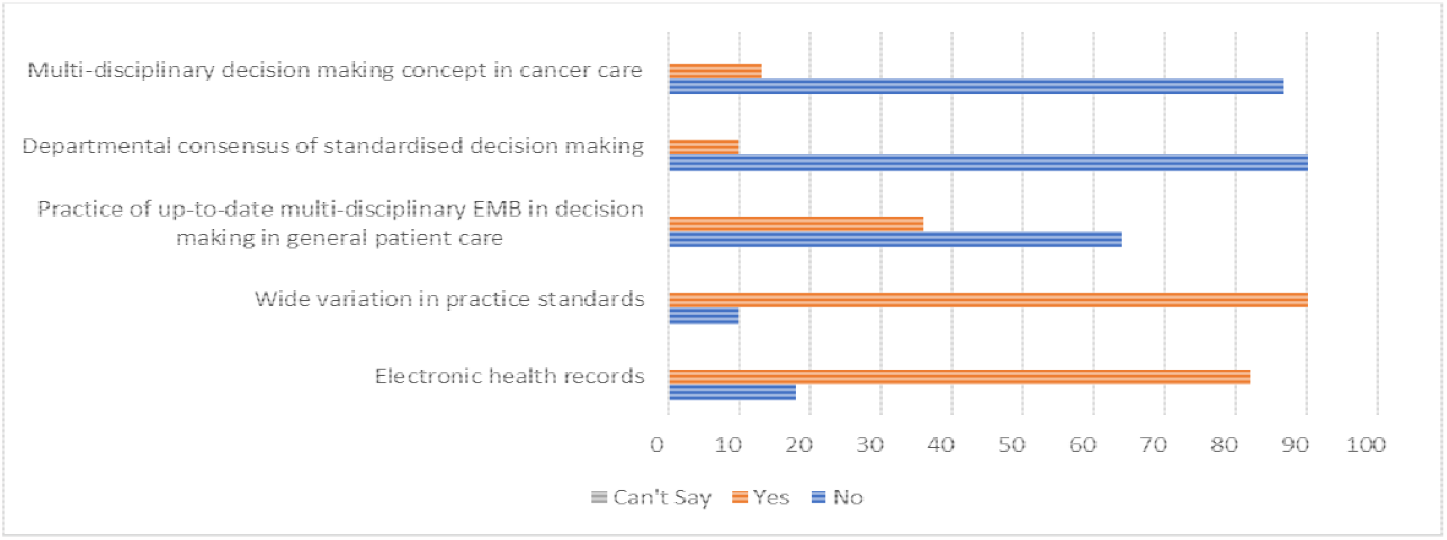
Percentage of participant answers on the favourability of current healthcare practises. **Summary of Fig. 2:** EHRs; 18% no, 82% yes, 0% can’t say. Wide variation in practice standards; 10% no, 90% yes, and 0% can’t say. The practice of up-to-date multi-disciplinary EBM in decision-making in general patient care; 63% no, 36% yes, 0% can’t say. The departmental consensus of standardised decision-making; 90% no, 10% yes, 0% can’t say. Multi-disciplinary decision-making concept in cancer care; 87% no, 13% yes, 0% can’t say.

82% of the study participants used EHRs in their clinical practises, which is the only positive attribute that emerged in this subsection, for the further digitalisation of healthcare with the incorporation of CDSS. However, the participants claimed to use basic versions of EHRs in their hospital settings.

Explaining the complexity of EHRs, one of the participants from a Tier 1 government hospital said, “the EHRs we use consist of scanned records of the written case sheets. Though it is theoretically an electronic record, it doesn’t offer the advantage of a conventional EHR by linking patient details to the results of the investigations or interlinking the results of diverse investigations like blood tests and CT^4^ scans. Hence, gathering patient information is sometimes easier on a paper-based system rather than electronic records.”

Another physician from a Tier 1 private institution explained that digitalisation is not the one-solution for everyone, “EHRs reflect well on the hospital but I serve 4 hospitals. With the need to attend to patients spread across 4 hospitals in a day, I can’t lose valuable time sitting at a computer desk. Therefore, I have set my practice to the paper-based system which is readily available and quite efficient. I use EHRs only as a backup.” Time constraints and the need to spend more time with the patients were also highlighted by another physician, “I need to ensure that I spend a good amount of time with my patients amongst my tight schedules. When there are 1-2 computers with people queuing up to access them, I cannot stand and wait. That is the time I should be spending with my patients.”

The lack of standardisation of care is evidenced by 90% of the study participants claiming wide variation of treatment, 90% acknowledging the absence of a diagnostic and research consensus, 87% denying the usage of SOPs^5^ in cancer care and a further 63% reporting the absence of SOPs in the decision-making process of current practises, is a notable roadblock.

One of the senior consultants explained how multidisciplinary decision processes are not formalised in India, “when I trained in the UK, multidisciplinary decision-making used to be a regular practice standard. However, the outpatient units here are always crammed up and we are required to work in a very hectic schedule with time constraints. So, we don’t have the luxury of dedicating a time slot for a multi-disciplinary meeting. Now, when I attend to a pre-diagnosed cancer patient, I call up the oncologist and share a snapshot of the patient record on WhatsApp and when he/she replies, I record it on the patient’s record. This is how the MDT^6^ system works here. While I am fully aware of CDSS through my experience in the Western countries, I don’t see any scope of its implementation in corporate practice here.”

Another senior consultant rejected the requirement of multidisciplinary decision-making by voicing out a personal belief, “I believe that the multi-disciplinary concept is ingrained in most clinicians and every doctor should develop that skill. In my practice, I’m responsible for all the therapeutic decisions for my patients. I decide about the requirement and the relevance of a diagnostic investigation, procedures, surgical interventions, chemotherapy, or endocrine therapy. I am the one-stop solution for all the requirements my patients have. Having said that, I cannot perform radiotherapy as I am not trained in that. I don’t see a place for CDSS in my practice as I’m personally equipped with these algorithms. For me, multi-disciplinary decision making is more of an academic exercise for teaching and training the trainees rather than a patient requirement.”

Addressing the lack of consensus and diverse practice standards, one physician said, “all the surgeons and physicians in our local area have an unwritten consensus on how cancer patients are to be treated. We are aware that the consultants have different preferences, so I see no point in holding an MDT meeting. As colleagues, we respect each other’s preferences and acknowledge that there are multiple ways of treating the same condition. At the end of the day, we are unified by our individual responsibilities to the patients and take decisions in their best regard. We offer our services and refer the patients to other specialists, if and when the need arises. In this climate, CDSS can’t add any value to the system unless we standardise our whole practice.” Another clinician further explained how a system that focuses on standardisation of care will not be beneficial in a sector where a wide variation of practice is the norm, “there are several right ways of treating the same condition. For my patients, I decide the treatment course and ultimately, it is down to the patient to accept or reject my suggestions. It is very likely that treatment practices vary among consultants. I’ll provide a suggestion to a colleague, only when I’m asked for one. CDSS will not work with the widely varying treatment options and therefore, for some, it might work as a wonderful system while for others it might be dismal. Hence, I think it can be widely adopted only if we can modify the system to cater to individual preferences.”

A consultant who recently joined a corporate facility reflected the same thought while questioned about the lack of institutional SOPs, “I am not aware of standard departmental practice or standard operating procedures. There is no such thing called departmental induction and I don’t see why that is necessary. The only department meeting we attend is a monthly morbidity and mortality meeting, where we discuss serious patient care issues like patient death. We have been practising medicine for a long time and therefore, we know the best practices. We are answerable only to the patient and to the court of law for practice standards. On CDSS, I would love to use it in my practice but I’m uncertain about how my colleagues will perceive it.”

### 3.2 Organisational and Infrastructural Concerns

While answering questions concerning the organisational and infrastructural readiness for newer innovative healthcare technology, the participants had more negative than positive responses, see Fig. 3.

**Fig. 3.**
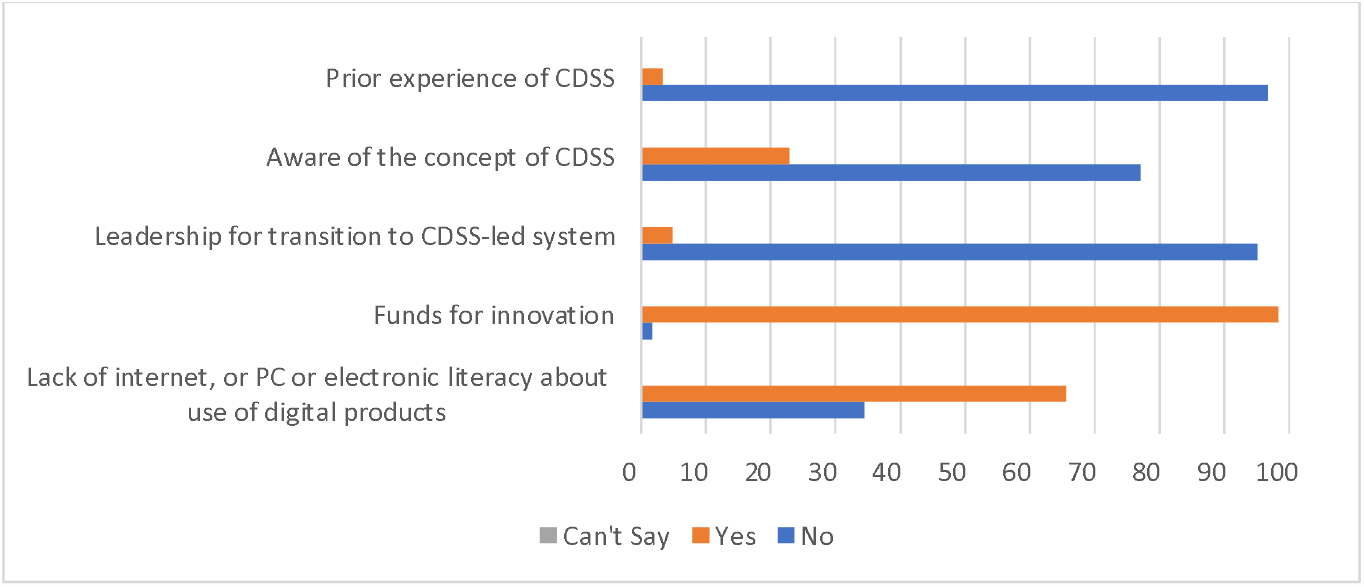
Percentage of participant answers on organisational and infrastructural issues regarding CDSS implementation. **Summary of Fig. 3:** Availability of internet, or PC or electronic literacy about use of digital products; 34% no, 66% yes, 0% can’t say. Funds for innovation; 2% no, 98% yes, 0% can’t say. Leadership for transition to CDSS-led system; 95% no, 5% yes, 0% can’t say. Aware of the concept of CDSS; 77% no, 23% yes, 0% can’t say. Prior experience of CDSS; 97% no, 3% yes, 0% can’t say.

98% of the participants confirming the organisational availability of funds dedicated to newer innovation and a further 66% claimed that their institutions had the necessary technical systems in place is a promising reflection of the infrastructural readiness. However, that is hampered by the manpower needed to efficiently work the systems. Only a mere 5% of the study population expressed enthusiasm to take charge and hold leadership positions necessary for CDSS incorporation if it was available free of charge. Around 3% of the experts had prior experience with CDSS systems, and a staggering 77% were unaware of the concept of CDSS.

Elaborating on the availability of funds for innovation, a C-suite level executive of Tier 1 corporate hospital said, “at our organisation, we have funds allocated for innovation. Usually, we provide these funds to start-ups with the potential for innovation. Recently we had given Indian rupee 5 lakhs for innovations. The start-ups produced certain results, but we are not sure how to promote it further. Though we go to start-up events and support innovation, any funds that we use for business problem solving will involve due diligence to ensure there is a good return on investment.”

A hospital administrator shed light on the lack of efficient manpower by discussing the techno-competence of senior physicians, “I’ve observed that the consultants aged 40 and above tend to possess poor computer literacy. Though they can work efficiently with their smartphones, their typing speed is generally not very good, so they require a computer-literate person (e.g. smart intern) to accompany them all the time. From the managerial perspective, it is difficult to find doctors who will work as transcribers, it is not the rewarding or challenging career they are after. Hence, the feasibility of the use of a system like CDSS becomes difficult, as that would require people to enter the data and generate results.”

### 3.3 Attitudes towards CDSS

Besides the aforementioned factors, attitudes towards a system will have a massive role in determining its success. Unfortunately, the attitudes towards CDSS were found to be not very favourable, see Fig 4.

**Fig. 4.**
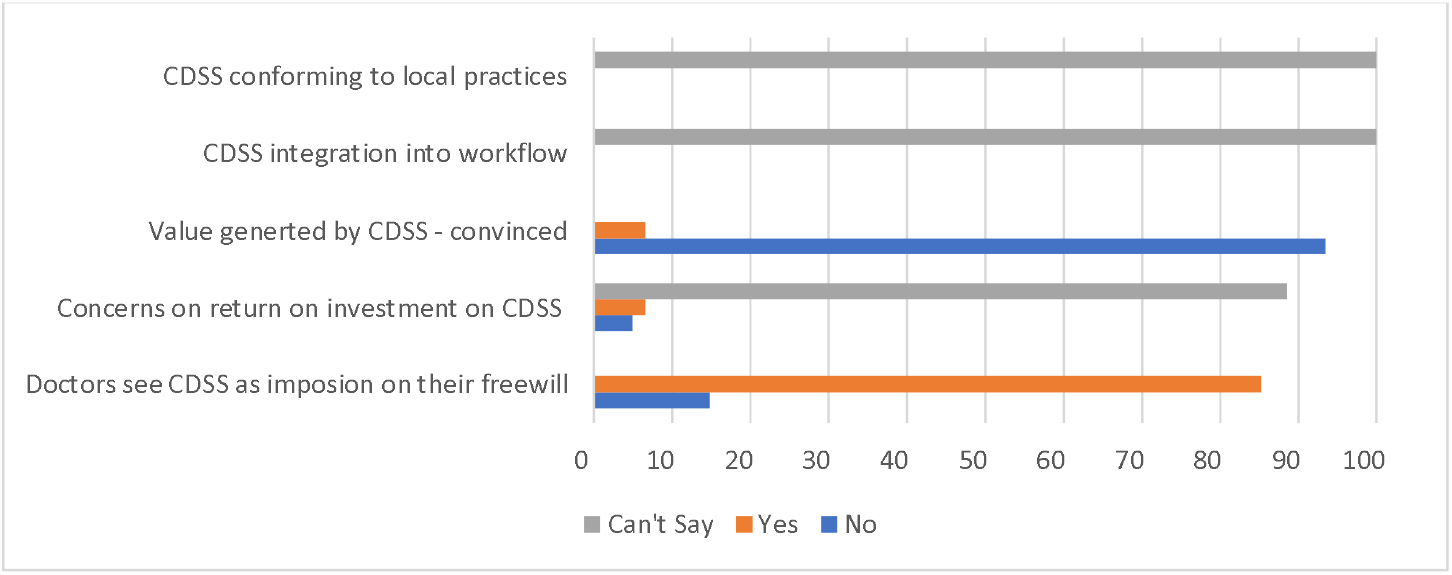
Percentage of participant answers on the user-adoption and attitudes towards CDSS. **Summary of Fig.4:** Doctors see CDSS as an imposition on their free will; 15% no, 85% yes, 0% can’t say. Concerns on return on investment on CDSS; 5% no, 7% yes, 88% can’t say. The value generated by CDSS; is 93% no, 7% yes, and 0% can’t say. Concerns on the integration of CDSS; 0% no, 0% yes, 100% can’t say. Concerns on CDSS conforming to local practises; 0% no, 0% yes, 100% can’t say.

While 85% of the population studied see CDSS as an imposition of their free will, a mere 7% were convinced of the value of integrating CDSS into the existing systems. A physician who saw CDSS as an imposition of decision-making freedom elaborated, “I would rather not use such a system in my practice as it would curb the freedom to make the best decisions for my patients. Clinical decisions should be taken jointly between the patient and the treating doctor. CDSS is not a party to this decision, even remotely. If we are to make it a standard practice, then I would be forced to make certain decisions without being convinced that is the right one.” Another doctor expressed fears over the system’s influence on clinical expertise, “why would I develop an unwanted dependence on a CDSS system? I would like to train my skills and decision-making skills. That will be the best investment as I am paid to make decisions. CDSS is not only unnecessary, but I’m also worried I might lose my skills if I use it in my practice.”

Talking about the value of CDSS in clinical practice, a consultant with experience with IBM Watson said, “I am one of the first few consultants to try IBM Watson in a multi-disciplinary decision-making process, but I figured out that it is no match to an expert. When I used the system in my practice, I found it to be largely time-consuming, and rather an academic exercise. Of course, if we have CDSS integrated into the EHRs, it might be useful in day-to-day practice but in the absence of widespread integration, I don’t think it can add any value to the practice.”

88% of the participants expressed uncertainty over the returns of introducing this healthcare innovation, while 5% were confident that it would not yield any return. When prompted to identify the issues surrounding the integration of CDSS and the conformation of CDSS to local practises, all of the participants failed to produce a response, which testifies to their uncertainty about CDSS.

Talking of the monetary concerns, a manager of a corporate hospital said, “One of the main issues we are facing is improving the flow of our patients and improving the flow of revenue to the hospital. I do not see how CDSS will be helpful to us. Our main income is from patient referrals, investigations, surgeries, and hospital stays. Anything which increases these will be considered as a good return on investment.”

A hospital administrator expressed concerns regarding the integration of CDSS into clinical workflow, “in our hospital, there are 2 groups of physicians and surgeons; one is the senior, digitally illiterate, experienced group while the other is the new generation of doctors who are digitally oriented and agile in technology adoption, but they are often side-lined by the powerful seniors. The senior group can be quite inflexible when it comes to technology and change in the model. In an institution where hierarchical medicine has a stronghold, it is difficult for me to imagine a smooth transition to digital health technologies such as CDSS.”

CDSS conformity to local practices was one of the integral issues identified, as one of the consultants said, “All the successful outcomes of IBM Watson are from Western studies. They are not applicable to the Indian subcontinent. For instance, we know that the breast cancer behaviour in the Afro-Caribbean population is significantly different to the Caucasian population and therefore, in the context of India, using an AI model framed in the Western set-up will not only be useless but can have unintended consequences too.”

### 3.4 Demographic Analysis

The results were further analysed to identify any patterns that emerged with regard to the demographic differences of the participants involved in the study; Tier 1 versus Tier 2 cities and state-run facility versus a corporate-run facility.

Comparing the responses between the participants from public and private organisations, a significant difference was observed with respect to the organisational and infrastructural concerns. Health practitioners in the private sector displayed 40% more awareness about CDSS when compared to their counterparts in the public sector. Furthermore, when compared to those in the public healthcare sector, the participants in the private setting were found to be 10% more enthusiastic to take initiative and lead teams for the successful incorporation of CDSS in their organisations. There were not any significant differences in the other aspects.

Wide variations were observed when the responses of participants from Tier 1 cities were compared with those of Tier 2 participants. 56% more participants from Tier 1 cities gave an account of the usage of institutional SOP in their current practises to facilitate decision-making. A similar pattern was observed with respect to the digitalisation of their workplaces – 49% of participants from Tier 1 institutions were more likely to have a digital workplace when compared to their Tier 2 counterparts, and 34% more clinicians in the Tier 1 cities had access to EHRs. The awareness of CDSS was more among participants in Tier 1 cities (44%). 28% more medical practitioners in Tier 2 cities compared to those in Tier 1 cities considered CDSS as an imposition on their will to make clinical decisions.

## 4.0 Discussion

The paradigm shift in today’s healthcare and complex clinical decision-making brought about by clinical decision support systems has resulted in widespread adoption (8,14,15,16,17,18). The electronic health record has shown a massive amount of workload automation and smart capabilities due to the integration of these advanced analytics systems. The enhanced safety, cost-efficiency, patient, and physician satisfaction have forced regulators, governments, and hospital owners to consider these smart systems.

The governments of the US, Canada, Denmark, Estonia, Australia and the United Kingdom endorse or incentivise the integration of clinical decision support systems into electronic health records (8,14). It has resulted in 41% of the US hospitals (15) and 62% of Canadian practitioners (16) integrating this technology into their workflow. CDSS-powered workflow has improved resource utilisation and has resulted in a cost savings of $717,538 per year without increasing the length of stay and mortality (21). In one study, CDSS was able to reduce 91.6% of medication consultations without introducing any errors (22).

The impact of CDSS has been reported to make a positive impact on laboratory resource utilisation (17), medication error prevention (22), accurate disease coding (23), improving the quality of medical records (20), improving the outcome following surgery (21,22), patient guidance (26), implementing best practice in patient services (23) and appropriate utilisation of radiological imaging (21). The diseases in which CDSS has shown evidence of clinical impact include but are limited to inflammatory bowel diseases, heart failure, hypertension, elderly care, sleep apnoea, diabetes, flu, electrocardiogram (ECG) analysis, arterial blood gas analysis, protein electrophoresis, blood cell counting, brain tumour classification and grading, bladder cancer grading and recurrence, Hepatitis-B and C testing, cardiac arrhythmia detection, tumour detection, medical imaging interpretation, diabetic retinopathy diagnosis, Alzheimer’s diagnosis and peripheral neuropathy (24,26,27,28,29,30,31,32,37,38,39,40).

Artificial Intelligence (AI) in healthcare is gaining momentum and popularity due to its immense potential to enhance the quality of care. However, the advances are largely limited to the western economies with the US and Europe holding 72% and 15% of the global CDSS market share, respectively (37). The incorporation of CDSS has huge potential to enable affordable health care delivery at a large scale and bridge the gaps in resource distribution in developing countries such as India (38,39,40). Developing economies deal with their unique array of challenges to achieve the seamless introduction of newer technology in their healthcare systems, due to its sheer population, complex sociocultural norms and vast demographic differences (41).

Due to the expanding population, vast talent pool, lack of standardisation of medical care and diverse population demographics, the Indian healthcare system poses a unique set of challenges and potentials (42). While there are various types of CDSS produced in the market, the usage will depend on several factors. Nevertheless, in India where 1 million cases of cancer are diagnosed annually (43), CDSS has the tremendous potential to improve the quality of cancer care with early detection and screening. Technology-enabled tuberculosis medication adherence systems can reduce the burden of Multidrug-Resistant Tuberculosis (MDR-TB) (44). Diabetic screening is another domain where CDSS can play a massive role, with the country predicted to emerge as the diabetic capital of the world (45). With machine learning algorithms, it is possible to bridge the gap and distribute scarce knowledge to the masses, in a bid to establish egalitarian healthcare. The current study was designed to assess the favourability of the Indian healthcare sector for the introduction of this digital healthcare technology.

Applying Hall’s Concerns-Based Adoption Model (13) to this study, 77% (n=47) of users had stage 0 or awareness concern at the outset where the survey participant was not aware if this was a concern as he/she has no idea about it while only 23% (n=14) had a higher level of concerns. When the potential users were educated about the CDSS, the number of people with level 1(information) & level 2 (personal) concerns increased to 85% (n=52). Here the users were more concerned about the threat arising out of the use of the system in their clinical practice e.g., the imposition of CDSS on their decision-making freewill. 93% (n=57) had raised a stage 3 or 4 concern e.g., CDSS might not cope with the existing logistics and systems in place, “largely time-consuming”, “the absence of widespread integration”, “an AI-model framed in the Western set-up” “can have unintended consequences too” etc. Most of the stage-4 concerns were related to the widespread scepticism surrounding the ROI and value generated by the integration of the CDSS. The collected data did not have any concerns about the collaborative, multidisciplinary workflow created by CDSS (e.g., level-5 concerns) or suggestions on ways and means to improve current models of CDSS available in the market (level-6). Earlier in this article, we observed the lack of standardised practice in India, evidenced by wide variation, lack of departmental consensus, multi-disciplinary decision-making, and standard operating procedures in cancer and general patient care. The vagaries of clinical care standards have endowed clinicians with a significantly higher degree of autonomy in decision-making.

It may prove to be a roadblock in the introduction of CDSS. The collation of all the interview data indicated that the perceived usefulness and perceived ease of use were big barriers to user adoption of this automation technology. This is a relative disadvantage as per Roger’s Diffusion of Innovations theory (46).

Moreover, the organisation’s readiness to embrace the innovation was marred by various infrastructural issues e.g., lack of reliability of internet, adequate number of computers and electronic literacy about the use of digital products. These infrastructural issues created a sense of misfit of digital technology in the eco-system, reducing the compatibility and increasing the complexity as per Roger’s Diffusion of Innovations (47). Even though in an academic setting good results can be demonstrated, integration issues reduced the real-world triability of the technology (47).

One of the most important factors for the successful adoption of new technology is observability (Roger’s Diffusion of Innovation) e.g., an easily noticeable result upon the use of the technology was lacking. Most hospital decisions on technology investments were based on return on investment such as saving of manhours, or generation of revenue. The technology can easily show its impact in settings where there are a high patient load compared to resources. However, in Tier 1 and 2 cities in India, there is a higher (almost double) density of doctors to patients and private practice is highly competitive.

Average socioeconomic status, poor digital literacy, less than average digital infrastructure and lack of technology leadership force the country to approach digital technologies such as CDSS with a high degree of scepticism, concerns and poor risk-taking. These factors are likely to leave the country as late majority or laggards. It is also possible that this technology may fail to diffuse due to a lack of awareness. This is in sharp contrast with the Western economies who are more open to the use of CDSS technologies. A study by French Breast oncologists showed that physician compliance with CDSS exceeded 90% when the technology provided the correct proposition (48). On the other hand, a study involving general practitioners in West Ireland showed that 94% of physicians were open to the use of CDSS despite 74% being unfamiliar with the technology (49).

In recent years, India has made substantial efforts to digitalise the existing healthcare system and make quality healthcare accessible to the marginalised sections of society. The Integrated Health Information Program (IHIP) is a testimony to the bold commitment shown by the nation to digitalise healthcare as it aims to provide an EHR for every citizen, integrate the existing EHRs to enable interoperability and establish uniform standards for EHRs across the country (50) In a bid to promote digitalisation of healthcare, the Ministry of Health and Family Welfare established National e-Health Authority (NeHA) in 2015 which aims to provide transparency in the maintenance and regulation of electronic transmissions of health information (51). A draft bill was also passed to ensure confidentiality of the digital clinical information, known as the Digital Information Security in Healthcare Act (DISHA) (42,53). The world’s largest government-funded health initiative, Ayushman Bharat introduced by the Government of India in 2018 caters to 50 crore people and the services are integrated with both private and public hospitals (53). These governmental initiatives are definite indicators of the country’s willingness and readiness to embrace newer innovations in the multi-layered healthcare sector.

The lack of product awareness and conviction appears to be two major barriers to the implementation of CDSS in Indian healthcare, where the market is in the stage of innovation and overcoming such barriers may be crucial to the adoption of the product into the market as other studies have shown (56,57). Studying the regional differences between Tier 1 and 2 cities, the hospitals in the former group may hold the key to bringing the technological transformation by being favourably placed to become agents of change by championing the ideas and values. Such a model use case will change the public attitude and increase user confidence (51). The high degree of usage of EHRs and availability of funds for innovation in hospitals was an encouraging trend as introducing advanced technology such as CDSS into a digitalised healthcare system is considerably easier.

## 5.0 Conclusions

India spends about 1.28% of its Gross Domestic Product (GDP) on its public healthcare (56). In comparison, many of the Organisation for Economic Co-operation and Development (OECD) countries spend between 9-16% of their GDP on the public healthcare system (58). Given the shocking level of patient safety issues amidst resource constraints and rising demand, Artificial Intelligence is the way forward. CDSS can have the greatest impact on the healthcare of developing economies by providing the highest level of healthcare standards while minimising healthcare expenditures. At the same time, it can also reduce medical errors and enhance patient safety by design and standardisation as discussed earlier.

Average socioeconomic status, poor digital literacy, less than average digital infrastructure and lack of technology leadership have forced India to approach digital technologies such as CDSS with a high degree of scepticism, concerns and poor risk-taking. These factors are likely to leave the country as late majority or laggards.

Despite all the impediments discussed earlier, India appears to be striving well to embrace digital health as seen by the many recent efforts e.g., National Digital Health IDs, Ayushman Bharat, the world’s largest insurance scheme and EHR for every citizen (59-62). Therefore, a high-quality CDSS cost-effectiveness study to showcase the benefit, safety, ease, and cost-saving potential of the use of CDSS will help the country to overcome the hesitancy associated with CDSS adoptions. The hospitals in Tier 1 cities appear to hold the key to bringing the technological transformation being favourably placed to become agents of change.

## Data Availability

All data produced in the present work are contained in the manuscript

## List of Abbreviations

AI: Artificial Intelligence
BIS: Bank for International Settlements
CDSS: Clinical Decision Support Systems
CT: Computed Tomography
DISHA: Information Security in Healthcare Act
ECG: Electrocardiogram
EHR: Electronic Health Record
GDP: Gross Domestic Product
Health IT: Health Information Technology
ID: Identity
IHIP: Integrated Health Information Program
MDR-TB: Multidrug-Resistant Tuberculosis
NeHA: National e-Health Authority
OECD: Organisation for Economic Co-operation and Development
WHO: World Health Organization
SOP: Standard Operating Procedure
MDT: Multi Disciplinary Team

## Declarations

### Ethics approval and consent to participate

At the outset, the research methodology was approved by YouDiagnose Ethical Approval Committee. This research work was commenced after this approval and this study did not involve any human data, tissues, samples, or materials. Consent to participate was obtained from each participant at the beginning of the study. Using the NHS Health Research Authority and United Kingdom Medical Research Council decision-making tool, it was determined that this study would produce generalisable or transferable findings. Therefore, informed consent was obtained from each participant before the structured interview. It was also determined that the study would anonymize the participants to mitigate any risk. All the methodologies were in line with the core practices of the Committee on Publication Ethics (COPE) and the Declaration of Helsinki.

### Consent for publication

Agree

### Availability of data and materials

The research data is shared with the readers and with the editorial board for the peer-review process. The datasets generated and/or analysed during the current study are not publicly available due to the databases being held anonymously but are available from the corresponding author at reasonable request. As per the terms of engagement, the participants held positions in various public and private institutes, and they have disallowed public disclosure of the institutes, participants’ names and positions. Many times, such disclosures might have repercussions in the office’s political circle. Many of the opinions may be misconstrued as tarnishing the reputation of institutions which might have consequences e.g., career progression etc.

### Authors’ contributions

N.M. and S.T. helped in designing the concept of the study. S.G. helped in arranging, conducting and collecting data for the interviews. A.M.1 and P.W. compiled and analysed the data. P.W., H.G. and N.K. were major contributors to manuscript writing. A.M. was a major contributor to each step of the current work. All authors read and approved the final manuscript.

## Footnotes

1 The World Health Organisation

2 Electronic Health Record

3 Market intelligence company

4 Computed Tomography scan used for diagnostic purposes in radiology

5 Standard Operating Procedure

6 Multi-Disciplinary Team

## References

1. World Health Organization. Health Systems: Improving Performance. 2000. https://apps.who.int/iris/bitstream/handle/10665/42281/WHR_2000-eng.pdf?sequence=1&isAllowed=y. Accessed 20 Dec 2021.

2. Healthcare in India: Private Vs. Public Healthcare. https://www.mapsofindia.com/my-india/india/healthcare-in-india-private-vs-public-healthcare. Accessed 20 Dec 2021.

3. Science, technology, and medicine in Colonial India. https://www.researchgate.net/publication/249096899_Science_Technology_and_Medicine_in_Colonial_India. Accessed 20 Dec 2021.

4. Business India. 2019. India’s doctor-patient ratio still behind WHO-prescribed 1:1,000: Govt.

5. The boom in digital healthcare is India’s opportunity to build global telemedicine companies. https://yourstory.com/2018/01/boom-digital-healthcare-indias-opportunity-build-global-telemedicine-companies/amp Accessed 20 Dec 2021.

6. Berner E., Tonya J, Le Lande T., 2007. Overview of Clinical Decision Support Systems. Clinical Decision Support Systems, 3–22.

7. Bajaj A., 2020. Healthcare industry in India. Invest India. https://www.investindia.gov.in/sector/healthcare. Accessed 20 Dec 2021.

8. Electronic Health Records: A Global Perspective. http://web.pdx.edu/~nwallace/GHS/EHRP2.pdf. Accessed 20 Dec 2021.

9. Praveen, D., Patel, A., McMahon, S. et al. A multifaceted strategy using mobile technology to assist rural primary healthcare doctors and frontline health workers in cardiovascular disease risk management: protocol for the SMARTHealth India cluster randomised controlled trial. Implementation Sci 8, 137 (2013). https://doi.org/10.1186/1748-5908-8-137

10. Malhotra, Savita et al. “Telepsychiatry clinical decision support system used by non-psychiatrists in remote areas: Validity & reliabilityof diagnostic module.” The Indian journal of medical research vol. 146, 2 (2017): 196–204. doi:10.4103/ijmr.IJMR_757_15

11. Praveen D, Patel A, Raghu A, Clifford G, Maulik P, Mohammad Abdul A, Mogulluru K, Tarassenko L, MacMahon S, Peiris D SMARTHealth India: Development and Field Evaluation of a Mobile Clinical Decision Support System for Cardiovascular Diseases in Rural India JMIR Mhealth Uhealth 2014;2(4):e54 URL: https://mhealth.jmir.org/2014/4/e54 DOI: 10.2196/mhealth.3568

12. Khairat S, Marc D, Crosby W, Al Sanousi A. Reasons For Physicians Not Adopting Clinical Decision Support Systems: Critical Analysis. JMIR Med Inform. 2018;6(2):e24. Published 2018 Apr 18. doi:10.2196/medinform.8912

13. Hall G., George A., Rutherford W., et al. Measuring stages of concern about the innovation: A manual for use of the SoC questionnaire. The University of Texas. 1977.

14. Nohr C., Parv L., Kink P., Cummings E., Almond H., Norgaard J., Turner P., 2017. Nationwide citizen access to their health data: analysing and comparing experiences in Denmark, Estonia and Australia. BMC Health Services Research. 2017;17:534.

15. Electronic Medical Record Adoption Model (EMRAM). https://www.himss.org/what-we-do-solutions/digital-health-transformation/maturity-models/electronic-medical-record-adoption-model-emram Accessed 20 Dec 2021.

16. Chang F., Gupta N., 2015. Progress in electronic medical record adoption in Canada. Canadian Family Physician. 2015;61:12.

17. Algaze C., Wood M., Pageler N., Sharek P., Longhurst C., Shin A. Use of a Checklist and Clinical Decision Support Tool Reduces Laboratory Use and Improves Cost. Pediatrics. 2016;137:1.

18. Pruszydlo M., Fritz S., Tichy T., Kaltschmidt J., Haefeli W. Development and evaluation of a computerised clinical decision support system for switching drugs at the interface between primary and tertiary care. BMC Medical Informatics and Decision Making. 2012;12:137.

19. Bell C., Jalali A., Mensah, E. A Decision Support Tool for Using an ICD-10 Anatomographer to Address Admission Coding Inaccuracies: A Commentary. Online Journal of Public Health Informatics. 2017;5(2):222.

20. Haberman S., Feldman J., Merhi Z., Markenson G., Cohen W., Minkoff H. Effect of clinical-decision support on documentation compliance in an electronic medical record. Obstetics Gynecology. 2009;114:311–317.

21. Turchin A., Shubina M., Gandhi T. NLP for patient safety: splenectomy and pneumovax. Proceedings of AMIA 2010 Annual Symposium, Washington DC.

22. McEvoy D., Gandhi T., Turchin A., Wright A. Enhancing problem list documentation in electronic health records using two methods: the example of prior splenectomy. BMJ Quality and Safety, 2018;27:1.

23. Rosenbloom S., Daniels T., Talbot T., McClain T., Hennes R., Stenner S., Muse S., Jirjis J., Jackson, G. Triaging patients at risk of influenza using a patient portal. National Library of Medicine, 19(4). Journal of the American Medical Informatics Association. 2012;19:4.

24. Georgiou A., Prgomet M., Markewycz A., Adams E., Westbrook J. The impact of computerized provider order entry systems on medical-imaging services: a systematic review. Journal of the American Informatics Association. 2011;18,3.

25. Blackmore C., Mecklenburg R., Kaplan, G., 2011. Effectiveness of clinical decision support in controlling inappropriate imaging. Journal of the American College of Radiology. 2011;8:1.

26. Benhamou P. Improving diabetes management with electronic health records and patients’ health records. Diabetes and Metabolism. 2011;37,4.

27. Kumar R., Goren N., Stark D., Wall D., Longhurst, C.. Automated integration of continuous glucose monitor data in the electronic health record using consumer technology. Journal of the American Medical Informatics Association. 2016;23,3.

28. Berner E. Clinical Decision Support Systems-Theory and Practice. 2nd ed. Berlin: Springer Science & Business Media; 2013.

29. Spyridonos P., Cavouras D., Ravazoula P., Nikiforidis G.. A computer-based diagnostic and prognostic system for asessing urinary bladder tumour grade and predicting cancer recurrence. Medical Informatics and the Internet of Medicine. 2002;27,2.

30. Tsolaki E., Svolos P., Kousi E., Kapsalaki E., Fezoulidis I., Fountas K., Theodorou K., Kappas C., Tsougos, I. Fast spectroscopic multiple analysis (FASMA) for brain tumor classification: a clinical decision support system utilizing multi-parametric 3T MR data. International Journal of Computer Assisted Radiology and Surgery. 2015;10,7.

31. Morkrid L., Rowe A., Elgstoen K., Olesen J., Ruijter G., Hall P., Tortorelli S., Schulze A., Kyriakopoulou L., Wamelink M., Van de Kamp J., Salomons G., Rinaldo, P. Continuous age- and sex-adjusted reference intervals of urinary markers for cerebral creatine deficiency syndromes: a novel approach to the definition of reference intervals. Clinical Chemistry. 2015;61,5.

32. Hannun A., Rajpurkar P., Haghpanahi M., Tison G., Bourn C., Turakhia M., Ng A. Cardiologist-level arrhythmia detection and classification in ambulatory electrocardiograms using a deep neural network. Nature Medicine. 2019;25,65–69.

33. Suzuki K., Chen, Y. Artificial Intelligence in Decision Support Systems for Diagnosis in Medical Imaging. 1st ed. Cham: Springer Nature; 2018.

34. IBM Watson for Oncology. https://www.ibm.com/watson-health/solutions/cancer-research-treatment. Accessed 20 Dec 2021.

35. Lunit Inc. https://www.lunit.io/en. Accessed 20 Dec 2021.

36. Gulshan V., Peng L., Coram, M. Development and Validation of a Deep Learning Algorithm for Detection of Diabetic Retinopathy in Retinal Fundus Photographs. Innovations in Health Care Delivery. 2016.

37. BIS Research Report. Clinical Decision Support Systems (CDSS) Market – Analysis and Forecast 2021-2030. https://bisresearch.com/industry-report/global-clinical-decision-support-systems-market.html. Accessed 20 Dec 2021.

38. Smadja N., Poda A., Ouedraogo A., Schmid J., Delory T., Le Bel J., Bouvet E., Lariven S., Jeanmougin P., Ahmad R., Lescure, F.. Paving the Way for the Implementation of a Decision Support System for Antibiotic Prescribing in Primary Care in West Africa: Preimplementation and Co-Design Workshop With Physicians. JMIR Publications. Journal of Medical Internet Research. 2020;22,7.

39. Dalaba M., Akweongo P., Williams J., Saronga H., Tonchev P., Sauerborn R., Mensah N., Blank A., Kaltschmidt J., Loukanova S.. Costs Associated with Implementation of Computer-Assisted Clinical Decision Support System for Antenatal and Delivery Care: Case Study of Kassena-Nankana District of Northern Ghana. Plos One, 2014; doi: https://doi.org/10.1371/journal.pone.0106416

40. Lewkowicz D., Wohlbrandt A., Boettinger E.. Economic impact of clinical decision support interventions based on electronic health records. BMC Health Services Research. 2020;20,871.

41. Urban health in India: many challenges, few solutions. https://www.thelancet.com/journals/langlo/article/PIIS2214-109X(15)00210-7/fulltext Accessed 20 Dec 2021.

42. India against Cancer. http://cancerindia.org.in/cancer-statistics. Accessed 20 Dec 2021.

43. Liu X., Blaschke T., Thomas B., Geest S., Jiang S., Gao Y., Li X., Buono E., Buchanan S., Zhang Z., Huang S. Usability of a Medication Event Reminder Monitor System (MERM) by Providers and Patients to Improve Adherence in the Management of Tuberculosis. International Journal of Environmental Research and Public Health, 14(10).

44. Atre S. The burden of diabetes in India. The Lancet Global Health, 2019;7,4.

45. Rao K., Peters D. Urban health in India: many challenges, few solutions. (online) The Lancet Global Health. 2015;3,12.

46. Eichner J., Das M. Challenges and Barriers to Clinical Decision Support (CDS) Design and Implementation Experienced in the Agency for Healthcare Research and Quality CDS Demonstrations. Agency for Healthcare Research and Quality. Agency for Healthcare Research and Quality. 2010.

47. Rogers E. Diffusion of Innovations. 5th ed. New York: Simon & Schuster; 2003.

48. Bouaud J., Spano J., Lefranc J., Zelek C., Jaulerry B., Zelek L., Durieux A., Tournigand C., Rousseau A., Vandenbussche P., Séroussi B. Physicians’ Attitudes Towards the Advice of a Guideline-Based Decision Support System: A Case Study With OncoDoc2 in the Management of Breast Cancer Patients. National Library of Medicine. Studies in Health Technology and Informatics. 2015;216,264–9.

49. Hor C., O’Donnell J., Murphy A., O’Brien T. General practitioners’ attitudes and preparedness towards Clinical Decision Support in e-Prescribing (CDS-eP) adoption in the West of Ireland: a cross sectional study. BMC Medical Informatics and Decision Making. 2010;10,2.

50. Integrated Health Information Program (IHIP). https://www.nhp.gov.in/integrated_health_information_program_mtl. Accessed 20 Dec 2021.

51. Electronic Health Record Standards for India. https://www.nhp.gov.in/her_standards_mtl_mtl. Accessed 20 Dec 2021.

52. DISHA: The First Step Towards Securing Patient Health Data In India. https://www.mondaq.com/india/healthcare/723960/disha-the-first-step-towards-securing-patient-health-data-in-india Accessed 20 Dec 2021.

53. Budget 2018: World’s largest govt-funded health care programme to benefit 10 cr families. https://timesofindia.indiatimes.com/business/india-business/budget-2018-worlds-largest-govt-funded-health-care-programme-to-benefit-10-cr-families/articleshow/62740312.cms. Accessed 20 Dec 2021.

54. Liebe, J D et al. “Investigating the roots of successful IT adoption processes – an empirical study exploring the shared awareness-knowledge of Directors of Nursing and Chief Information Officers.” BMC medical informatics and decision making vol. 16 10. 27 Jan. 2016, doi:10.1186/s12911-016-0244-0

55. Moxham C, Chambers N, Girling J, Garg S, Jelfs E, Bremner J. Perspectives on the enablers of e-heath adoption: an international interview study of leading practitioners. Health Serv Manage Res. 2012 Aug;25(3):129–37. Doi: 10.1258/hsmr.2012.012018. PMID: 23135887.

56. Estimated value of public health expenditure in India from financial year 2017 to 2020. https://www.statista.com/statistics/684924/india-public-health-expenditure/. Accessed 20 Dec 2021.

57. Current health expenditure (% of GDP).: https://data.worldbank.org/indicator/SH.XPD.CHEX.GD.ZS. Accessed 20 Dec 2021.

58. Esmaeilzadeh P., Sambasivan M., Kumar N., Nezakati H. Adoption of clinical decision support systems in a developing country: Antecedents and outcomes of physician’s threat to perceived professional autonomy. International Journal of Medical Informatics. 2015;84,8.

59. India launches digital health IDs. https://healthcareglobal.com/digital-healthcare/india-launches-digital-health-ids Accessed 20 Dec 2021.

60. Electronic Health Record for Every Citizen. https://ehealth.eletsonline.com/2021/02/electronic-health-record-for-every-citizen/. Accessed 20 Dec 2021.

61. World’s Largest Insurance Scheme Ayushman Bharat Launches Mobile App. https://inc42.com/buzz/ayushman-bharats-pm-jay-app-launched-records-10k-installs/ Accessed 20 Dec 2021.

62. National Health Authority. https://pmjay.gov.in/about/pmjay. Accessed 20 Dec 2021.

